# Population density and basic reproductive number of COVID-19 across United States counties

**DOI:** 10.1101/2020.06.12.20130021

**Authors:** Karla Therese L. Sy, Laura F. White, Brooke Nichols

## Abstract

The basic reproductive number (R_0_) is a function of contact rates among individuals, transmission probability, and duration of infectiousness. We sought to determine the association between population density and R_0_ of SARS-CoV-2 across U.S. counties, and whether population density could be used as a proxy for contact rates. We conducted a cross-sectional analysis using linear mixed models with random intercept and fixed slopes to assess the association of population density and R_0_. We also assessed whether this association was differential across county-level main mode of transportation-to-work percentage. Counties with greater population density have greater rates of transmission of SARS-CoV-2, likely due to increased contact rates in areas with greater density. The effect of population density and R_0_ was not modified by private transportation use. Differential R_0_ by population density can assist in more accurate predictions of the rate of spread of SARS-CoV-2 in areas that do not yet have active cases.

**Article Summary Line:** U.S. counties with greater population density have greater rates of transmission of SARS-CoV-2, likely due to increased contact rates in areas with greater density.

## Introduction

The COVID-19 pandemic has infected millions of people globally, and there are over 400 thousand reported deaths and 7 million confirmed cases of COVID-19 worldwide.^1^ Transmission of airborne and directly transmitted pathogens,^2-4^ such as SARS-CoV-2 (the causative agent of COVID-19), have been previously shown to be density-dependent. Population density facilitates transmission of disease via close person-to-person contact,^5-8^ and may support sustained disease transmission due to increased contact rates.^9-11^ Large urban areas have more opportunities for disease transmission, and hotspots of SARS-CoV-2 have been mostly concentrated in cities.^12^

The basic reproductive number (R_0_) describes the contagiousness and transmissibility of pathogens, and is a function of contact rates among individuals, transmission probability, and number of infective individuals.^13^ Thus, R_0_ estimates of COVID-19 are not exclusively determined by the pathogen, and variability in R_0_ depends on local sociobehavioral and environmental settings, including population density.^12^ During the initial phase of the outbreak, or the exponential growth period, we hypothesize that spatial heterogeneity in R_0_ occurs in part due to geographic variability in contact rates, since transmission probability and population size remain constant. Since the exponential growth period occurs prior to the implementation of non-pharmaceutical interventions (NPIs), such as face coverings and social distancing, we would expect that the probability of transmission per contact would be the same equal across settings. Moreover, contact networks are also affected by transportation systems that facilitate disease spread due to increased interconnectivity and mobility between different geographic areas;^14,15^ thus, we also hypothesize that the association of population density and R_0_ may be differential depending on transportation accessibility, and areas that lack access to efficient modes of transportation would not have the same SARS-CoV-2 growth rate, even in high density areas.

In the current COVID-19 pandemic, data-driven analyses of the association of population density and disease transmission has not been systematically quantified. The estimation of differential R_0_ using these area-level factors can assist in more accurate predictions of the rate of spread of SARS-CoV-2 in geographic settings where cases have only begun to rise. In this study, we examine the association of population density with R_0_ of COVID-19 across United States counties.

## Methods

### Data

We obtained publicly available daily COVID-19 case and death data among United States counties from the New York Times.^16^ For each county, we assumed that the exponential growth period was one week prior to the second daily increase in cases. We assumed that the period of exponential growth approximately lasted 18 days, as previous research have shown the COVID-19 exponential period to be around 20-24 days in New York City,^17^ and calibrated it accordingly to create reasonable curves that approximated exponential growth across the counties (**Supplemental Materials Appendix 1**). The algorithm ensured that the virus had taken hold in the area and allowed a sufficient number of days to estimate the exponential growth rate. We restricted calculation of R_0_ to counties with greater than 25 cases at the end of the exponential growth period, as R_0_ cannot be estimated accurately with sparse data and it would be uncertain if the county was experiencing a sustained outbreak with community transmission. Data on the primary mode of transportation to work and median household income were obtained from the most recent 5-year American Community Survey (ACS) 2014-2018 survey estimates from the United States Census Bureau.^18^ Population and land area were obtained from the 2010 census, and density was calculated by population divided by total square km. All census data were extracted using the R package *tidycensus*.^19^

### Statistical analyses

We first compared the densities of counties included in the final analytical sample to those that did not have sufficient case counts with a two-sample Wilcoxon test. R_0_ was calculated using the Wallinga and Lipsitch method.^20^ We then conducted a cross-sectional analysis using linear mixed models with random intercept for each county and fixed slopes to assess the association of population density and R_0_. The models controlled for state-level effects using random intercepts, and county-level main mode of transportation to work percentage. We also adjusted for median household income to control for any potential confounding between the association of private car ownership and R_0_.

We fit 4 models with R_0_ as the outcome and the following factors as covariates: Model 1: population density; Model 2: population density and the percent of individuals reporting private transportation as their main mode of transportation to work; Model 3: population density and the percent of individuals reporting private transportation, and median household income; Model 4: population density, percent of individuals reporting private transportation, median household income, and the interaction of private transportation use with population density.

### Sensitivity analyses

We conducted three sensitivity analyses to address the limitations of our approach and assess the robustness of our results. First, we conducted the main analysis using death counts to estimate R_0_ to limit bias due to differential availability of testing by geographic location. We used the same exponential period as the cases, but with a lag of 14 days to account for the delays from symptom onset to deaths of cases.^21,22^ Moreover, the analysis of deaths was restricted to counties with greater than 10 deaths and more than 5 daily increases in incident deaths, in order to appropriately estimate R_0_ in counties with sufficient death counts. Second, we excluded counties within a radius of 15 miles, the average commuting miles in the United States,^23^ from counties with densities greater than the 75^th^ percentile. Removing these adjacent counties would demonstrate the extent of biases due to individuals commuting from surrounding counties to cities. If cases are imported from more densely population (i.e. cities) to less dense counties, we could potentially be biasing our estimates downwards. Lastly, we conducted an analysis excluding influential counties with a Cook’s distance measure over 4/N for each model, in order to ensure that findings were not driven by influential data points.

All analyses was conducted in R version 4.0.0.^24^ The figure and removal of adjacent counties in the sensitivity analyses were done with ArcGIS.^25^

## Results

The United States has 3,221 counties and county equivalents. When restricting to counties with greater than 25 cases, 1,151 (35.73%) counties were included (**Figure 1**). The median density in counties included and not included in the analysis were 59.8 people/km^2^ (IQR: 23.64-150.47) and 11.2 people/km^2^ (IQR: 3.65-23.64) respectively, and the difference was statistically significant (p <0.0001). The median R_0_ among the counties was 1.66 (IQR: 1.34-2.11).

**Figure 1.**
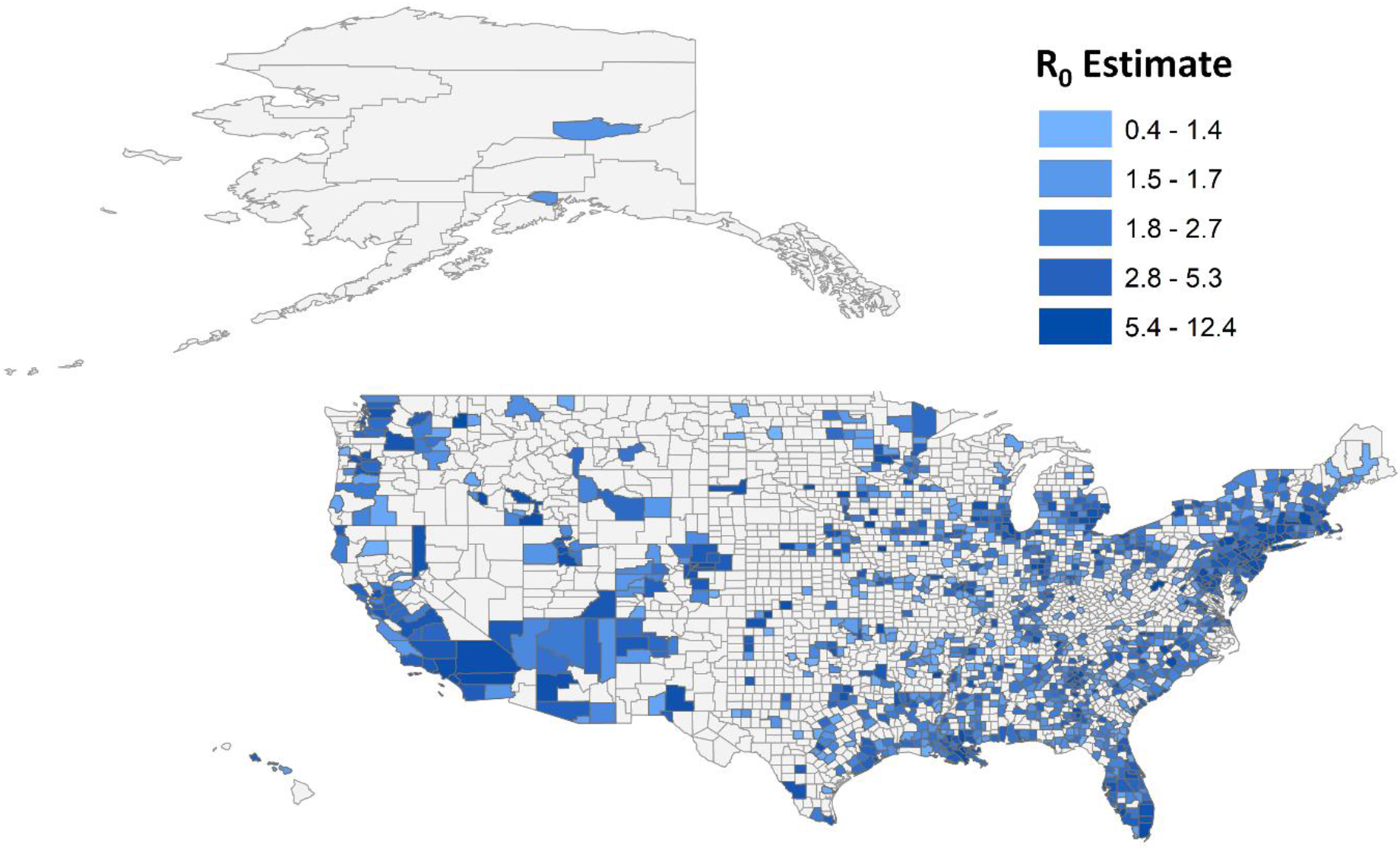
Basic reproductive number (R_0_) estimates across United States counties. Larger R_0_ indicates greater transmission during the initial phase of the outbreak, or the exponential growth period. We restricted calculation of R_0_ to counties with greater than 25 cases at the end of the exponential growth period (n=1,151), as R_0_ cannot be estimated accurately with sparse data and it would be uncertain if the county was experiencing a sustained outbreak with community transmission.

An increase in one unit of log population density increased R_0_ by 0.16 (95% CI=0.13 to 0.19) (Model 1; **Table 1**), or the doubling of population density increased the R_0_ on average by 0.11 (95% CI=0.09 to 0.13). When adjusted for percent of private transportation and median household income, the association of log population density and R_0_ remained unchanged (Model 3; **Table 1**).There was no significant interaction, and the effect of population density on R_0_ was the same among counties with a larger percentage with private cars as their transportation to work (Model 4; **Table 1**). R_0_ decreased by 0.12 (95% CI= -0.02 to -0.04) with an 10% increase in private transportation as the main commute mode, accounting for population density and median household income (Model 3; **Table 1**).

**Table 1.** Linear mixed models (random intercept, fixed slope) evaluating the association between population density and basic reproductive number (R_0_) among United States counties. Estimates for each model is a slope (beta) with a null of 0; a positive slope indicates that an increase in the log of population density increases R_0_ by the beta estimate for the log of population density. The interaction term indicates that the association of population density and R_0_ differs depending on the percentage of people using the private transportation for work.

|  | **Model 1** | **Model 2** | **Model 3** | **Model 4** |
| --- | --- | --- | --- | --- |
| Log of population density | **0.16\*\*\***<br>(0.13-0.19) | **0.15\*\*\***<br>(0.12, 0.18) | **0.15\*\*\***<br>(0.12, 0.18) | **0.2\***<br>(-0.06, 0.47) |
| Percent of private transportation (10%) |  | **-0.12\*\*\***<br>(-0.19, -0.04) | **-0.12\*\***<br>(-0.2, -0.04) | -0.08<br>(-0.27, 0.11) |
| Median household income (\$10,000) | | | 0<br>(-0.03, 0.03) | 0<br>(-0.03, 0.03) |
| Interaction of population density and transportation |  |  |  | -0.01<br>(-0.04, 0.02) |

pvalue \* < 0.05; \*\* < 0.01; \*\*\* < 0.001
Model 1- unadjusted association of population density and $R_0$
Model 2 – association of population density adjusted for the percent of individuals reporting private transportation
Model 3 – association of population density, percent of individuals reporting private transportation, and median household income
Model 4 – association of population density, percent of individuals reporting private transportation, median household income, and the interaction of population density and transportation

In all three sensitivity analyses, population density remained positively associated with R_0_, demonstrating the robustness of our main analysis. First, death data was used to calculate R_0_ from 310 counties. The median R_0_ among the counties that had sufficient death counts was 1.40 (IQR: 1.05-1.78). The unadjusted association between population density and R_0_ remained consistent (β=0.18, 95% CI=0.14 to 0.23) (Model 1a; **Table 2**), and there were no significant interactions (Model 4a; **Table 2**). Next, there were 288 counties above the 75^th^ percentile, and 414 counties that were within 15 miles of these counties high-density counties. We removed these 414 counties from the sample, and using the subsample of 737 counties, our findings remained consistent (**Table 2**). Influential counties were also not driving the association of population density and R_0_, and our results remained robust (**Table 2**). For the two sensitivity analyses excluding counties adjacent to highly dense counties and excluding high influence counties, however, the association of private transportation usage and R_0_ did not remain (Models 2a, 3a, 2b, 3b; **Table 2**).

**Table 2.** **Sensitivity analysis of linear mixed models (random intercept, fixed slope) using (a) deaths only, (b) removing counties within 15 miles of high density counties, and (c) removing high influence counties**

|  | Model 1 | Model 2 | Model 3 | Model 4 |
| --- | --- | --- | --- | --- |
| **Deaths only** |  |  |  |  |
| Log of population density | **0.18\*\*\***<br>(0.14, 0.23) | **0.14\*\*\***<br>(0.09, 0.19) | **0.12\*\*\***<br>(0.07, 0.17) | **0.48\***<br>(0.01, 0.94) |
| Percent of private transportation (10%) |  | **-0.16\*\***<br>(-0.26, -0.06) | **-0.15\*\***<br>(-0.24, -0.05) | 0.14<br>(-0.24, 0.53) |
| Median household income (\$10,000) | | | **0.06\*\***<br>(0.03, 0.1) | **0.07\*\*\***<br>(0.03, 0.11) |
| Interaction of population density and transportation |  |  |  | -0.04<br>(-0.09, 0.01) |
| **No counties adjacent to high density counties** |  |  |  |  |
| Log of population density | **0.17\*\*\***<br>(0.14, 0.2) | **0.17\*\*\***<br>(0.13, 0.2) | **0.16\*\*\***<br>(0.13, 0.2) | 0.02<br>(-0.13, 0.17) |
| Percent of private transportation (10%) |  | 0.01<br>(-0.07, 0.1) | -0.01<br>(-0.12, 0.1) | -0.12<br>(-0.28, 0.03) |
| Median household income (\$10,000) | | | 0.02<br>(-0.03, 0.06) | 0.01<br>(-0.03, 0.05) |
| Interaction of population density and transportation |  |  |  | 0.03<br>(0, 0.06) |
| **Removing high influence counties** |  |  |  |  |
| Log of population density | **0.18\*\*\***<br>(0.15, 0.2) | **0.18\*\*\***<br>(0.16, 0.21) | **0.18\*\*\***<br>(0.15, 0.2) | **0.38\***<br>(0.04, 0.72) |
| Percent of private transportation (10%) |  | -0.01<br>(-0.09, 0.07) | -0.02<br>(-0.1, 0.07) | 0.09<br>(-0.1, 0.29) |
| Median household income (\$10,000) | | | 0<br>(-0.03, 0.02) | 0<br>(-0.02, 0.03) |
| Interaction of population density and transportation |  |  |  | -0.02<br>(-0.06, 0.02) |

pvalue \* < 0.05; \*\* < 0.01; \*\*\* < 0.001
Model 1- unadjusted association of population density and R0
Model 2 – association of population density adjusted for the percent of individuals reporting private transportation
Model 3 – association of population density, percent of individuals reporting private transportation, and median household income

## Discussion

Our findings show that the basic reproductive number (R_0_) is associated with population density, even when percent of individuals that use private transportation and median income were accounted for. In these settings, greater population density may potentially facilitate interactions between susceptible and infectious individuals in densely-population networks, which sustain continued transmission and spread of COVID-19. Moreover, we see that population density continues to have an important impact on disease transmission regardless of transportation accessibility and median income, suggesting that the opportunity for effective contacts are mostly driven by crowding in denser areas, increasing the contact rates necessary for disease spread. However, we did not see that density-dependence is differential across transportation accessibility. Even though transmission is less in lower density areas (i.e. rural areas), rural settings may eventually disproportionately be more vulnerable to COVID-19 morbidity and mortality. Individuals in rural areas are generally older, have more underlying conditions, have less access to care, and have fewer ICU beds, ventilators, and facilities needed for severe COVID-19 treatment.^26-28^ Further research is needed on the overall burden of COVID-19 across the spectrum of population density.

Geographic estimates of R_0_ of SARS-CoV-2 need to take into account the specific area’s population density, since the R_0_ estimate is dependent on both the pathogenicity of the virus as well as environmental influences. In countries where cases are only on the starting to climb, such as countries in Latin America and Southern Africa,^1,29^ or there is a resurgence of cases, such as India, Iraq, and Israel,^30^ area-specific density can assist in predictions of R_0_, which is important because epidemiological forecasts and predictive models are sensitive to small changes in R_0_ inputs. Accurate estimation of R_0_ consequently lead to more precise estimates of the epidemic size, so that governments can appropriately allocate resources and coordinate mitigation strategies. Moreover, as cities and states reopen in the United States, and if there is a second-wave of infections, areas with higher density accessibility will likely have greater SARS-CoV-2 resurgence.

Our study has a number of limitations. While we demonstrate that population density is associated with R_0_, we estimated R_0_ based on the number of reported cases; therefore, the incidence of COVID-19 across US counties may be underestimated at varying rates due to differential testing. Testing data at the county-level currently do not exist, and we were unable to adjust for the number of tests performed. To mitigate this limitation, we included a random intercept term to adjust for state-level effects, and thus differential testing across states were accounted by our model. Differential testing by local governments within states are less likely to strongly impact our findings, as most funding and budgets for COVID-19 is distributed at the state-level.^31,32^ We also conducted a sensitivity analysis using death data which demonstrates the robustness of our findings. Additionally, we had to limit our analysis to counties that had sufficient case data in order to accurately estimate R_0_. Given our findings that the counties excluded in the analysis had a significantly lower density and presumably very low R_0_ due to lack of cases, the true association between population density and R_0_ would likely be greater than what we report in our analysis. Another limitation is that our model also assumes homogenous mixing, which may can be an oversimplification of the heterogeneity in contact patterns within populations.^4,33^ However, previous research has shown that population structure only changes R_0_ estimates slightly,^34^ and assumptions of well-mixed populations are valid in small-to-medium spatial scales.^35^ Moreover, our method loses spatial granularity in assessing R_0_ in counties, especially in counties with spatially heterogenous clustering. The aim of our study, however, was to provide a generalizable estimate of the association between population density and R_0_, in order to appropriately estimate potential for disease transmission, rather than a microspatial estimate that may not be generalizable to other settings. Finally, an important confounder that we were unable to adjust for is the number of importations of SARS-CoV-2 in these counties, as more urbanized areas are more likely to have links with countries and other states where the virus could have originated from. Even so, we still see that once an area is seeded with COVID-19, the growth rate is greater in denser areas during the time period prior to implementation of NPIs.

In summary, counties with greater population density have greater rates of transmission of SARS-CoV-2, likely due to increased contact rates in areas with greater population density. Population density affects the network of contacts necessary for disease transmission, and SARS-CoV-2 R_0_ estimates need to consider this variability for proper planning and resource allocation, particularly as epidemics newly emerge and old epidemics resurge.

## Data Availability

Daily COVID-19 case and death data among United States counties from the New York Times are publicly available.

https://github.com/nytimes/covid-19-data/blob/master/us-counties.csv

## Author contributions

KTLS, BEN contributed to conceptualization. KTLS contributed to data acquisition. KTLS, LFW, and BEN contributed to data analysis. All authors contributed to interpretation of results and manuscript writing.

## Disclaimers

The author’s views expressed in this publication do not necessarily reflect the views of the United States Agency for International Development or the United States Government.

